# Wastewater SARS-CoV-2 and COVID-19 Hospital Admission and Mortality – A Controlled Experimental Study

**DOI:** 10.64898/2025.12.19.25342702

**Authors:** Yalda Zarnegarnia, Samantha Abelson, Johnathon Penso, Kristina Babler, Mark Sharkey, Mario Stevenson, George Grills, Christopher E. Mason, Helena Solo-Gabriele, Erin Kobetz, Yan Guo, Naresh Kumar

## Abstract

After the COVID-19 pandemic, wastewater monitoring is increasingly used for infectious disease surveillance. Using the data from a controlled experimental hospital setting, this paper examines the association wastewater SARS-CoV-2 with COVID-19 hospital admission and mortality, and whether this association varies by patients’ characteristics. Weekly wastewater samples were collected from the University of Miami (UM) hospitals where COVID-19 patients were admitted from February 2020 to October 2022, and SARS-CoV-2 was quantified using qPCR. Data on hospital admissions and their mortality and demographic characteristics and comorbidities were acquired from the UM hospitals. Using factor analysis and hierarchical clustering, patients were stratified into four clusters. Frist, we examined cross-correlations between time-lagged COVID-19 hospital admission and mortality, and time-lagged SARS-CoV-2 to identify appropriate time-lags. Second, we modelled daily hospital COVID-19 cases and mortality with respect to time-lagged SARS-CoV-2, vaccine status and time-lagged COVID-19 hospital cases (as proxy of the risk factor for the transmission of the disease for each cluster separately and for all clusters together. 1,856 COVID-19 patients were admitted in the UM hospitals during the study period and 347 (18.7%) of them died. In cluster 4 that represented patients with preexisting chronic health conditions and intubation, the fatality rate was 59%. COVID-19 hospital admission showed strong (temporal) autocorrelation, suggesting that the preexisting cases can indicate the transmission rate of infection. Our analysis suggests that a 1% increase in SARS-CoV-2 was associated with a 0.28% increase in COVID-19 related hospital admission (β ∼ 0.275; 95 % CI = 0.18 to 0.37; p < 0.01). Both a week lagged auto-regressive COVID-19 cases and SARS-CoV-2 in wastewater together explained 89% of the total variation in hospital admission due to COVID-19. Among four clusters, the second cluster of minority communities showed the strongest association between time-lagged SARS-CoV-2 in wastewater and hospital admissions due to COVID-19 followed by cluster 1 of adult patients with low prevalence of preexisting health conditions. However, time-lagged wastewater SARS-CoV-2 did not show any significant association with COVID-19 hospital admission for patients with the pre-existing health conditions. A week lagged wastewater SARS-CoV-2 did not show any significant association with COVID-19 mortality. Our results indicate that the association between time-lagged wastewater SARS-CoV-2 and COVID-19 hospital admission varied by patients’ characteristics, suggesting variations in SARS-CoV-2 shedding by patients’ characteristics. These findings warrant to incorporate patient-specific demographic characteristics and comorbidities in modelling infectious diseases surveillance using wastewater monitoring of the infectious agents.

## 1. INTRODUCTION

The severe acute respiratory syndrome coronavirus-2 (SARS-CoV-2) quantitation in wastewater (WW-SARS-CoV-2 here after) is increasingly recognized as a useful method for the surveillance of COVID-19.^1-4^ It offers several advantages over the traditional clinical and serological surveillance methods, because it is non-invasive and cost-effective, and it captures the infectious agents shed (into wastewater) by both (a)symptomatic individuals many days prior to their clinical diagnosis. Emerging literature suggests that the WW-SARS-CoV-2 can predict COVID-19 incidence many days prior to their clinical diagnosis across different communities.^5^ However, there are inconsistencies in the performance of WW-SARS-CoV-2 for predicting COVID-19 cases.^6-8^ A host of factors can influence this performance.^9^ First, most studies use aggregated data, such as by county or state to develop COVID-19 prediction model using WW-SARS-CoV-2. Aggregation of these data at coarser spatiotemporal resolutions, such as by county and week, can introduce uncertainty and/or bias. For example, WW-SARS-CoV-2 for a given wastewater treatment plant may not fully capture SARS-CoV-2 shed by all (a)symptomatic cases in the county, and many homes in a jurisdiction may not be connected to that sewer plant.

Second, population demographics, local- and regional policies, community perception of the risk, vaccination, pre-existing health conditions, viral strain and viral shedding rate can also influence the relationship between WW-SARS-CoV-2 and COVID-19.^9,10^ For example, viral load shed by vaccinated individuals can vary from those unvaccinated. Age (65+), body mass index (BMI) ≥40, pre-existing cardiovascular diseases, hypertension, and behavioral choices, such as smoking, can also increase the severity of the disease.^11^ Moreover, there are disparities in COVID-19 hospitalization rate by race/ethnicity.^12-21^ Low-income minority populations were at a greater risk of contracting COVID-19, and COVID-19 related hospital admission and morality.^22^

Third, most studies rely on publicly available COVID-19 incidence data, which may not fully represent the severity of the disease measured by hospital admission and fatality rate. If an infection does not cause severe symptoms, it may affect people’s behavior, including proactive screening or testing, which can greatly affect the reported number of cases and clinical screening for the disease.

To address the above research gaps, we use patient-specific COVID-19 and WW-SARS-CoV-2 data from a hospital to pursue two aims: a) examine whether the relationship between WW-SARS-CoV-2 and COVID-19 cases in the hospital varies by patient characteristics and explore the efficiency of WW-SARS-CoV-2 to predict of COVID-19 hospital admission and mortality adjusted by patients’ characteristics.

## 2. MATERIALS AND METHODS

For this research we used multiple data sets. Weekly wastewater samples were collected from two manholes of the University of Miami (UM) hospitals from 9/30/2000 to 10/04/2022. Samples were concentrated by electronegative filtratration and lysed in DNA/RNA Shield (Zymo). RNA was extracted from the samples (Quick-RNA^TM^ Viral Kit, Zymo Research) and SARS-CoV-2 N3 gene was quantified using qPCR using protocols described.^8, 23^ Quality control measurements indicated that sample recoveries were 20% on average and that sample amplification inhibition was negligible. Deidentified patients’ data were acquired from the UM hospital for admission for COVID-19, which included date of admission, patients’ socio-demographic characteristics, health insurance status, pre-existing medical conditions, and medical conditions, such as ICU admission, ventilator use for the study duration. This data was provided following approved human subjects protocols (University of Miami Internal Review Board, IRB 20210164). The vaccination data for the Miami-Dade County were acquired from the CDC.^24^

Factor analysis of mixed data (FAMD), as a mixed method between principal component analysis (PCA) and multiple correspondence analysis (MCA), was used to examine similarities in different characteristics of patients. We then used hierarchical clustering on two the factor scores of the top two factors to partition data into four clusters. Weekly WW-SARS-CoV-2 data from two sites were averaged, resulting in weekly WW-SARS-CoV-2. Although the temporal scale of COVID-19 patient data was daily, these data for each cluster were also aggregated by week to match with the temporal resolution of WW-SARS-CoV-2 data. Time-lagged WW-SARS-CoV-2 were computed. WW-SARS-CoV-2 data were log-transformed, because these data were skewed. In the exploratory analysis, cross correlation function (CCF) was used to identify the relationship between time-lagged WW-SARS-CoV-2 for COVID-19 cases, which guided our final regression model. We used log-log regression to model COVID-19 cases with respect to four week lagged autocorrelation weighted WW-SARS-CoV-2 ((lag1*autocorrelation + lag2*autocorrelation, …, lag4*autocorrelation) / sum of autocorrelation). The goodness of fit, ACF and PACF of the residuals were explored. Analyses were conducted for each cluster separately and for all clusters together. Similarly, we modelled COVID-19 mortality with respect to time-lagged WW-SARS-CoV-2 adjusting for the vaccination rates.

## 3. RESULTS

A total of 1,856 patients were admitted to the UM hospitals for COVID-19 infection during the study period from 9/30/2020 to 10/04/2022. Patients’ characteristics are summarized in Table 1; 67.9% and 63.1% of the patients identified themselves as White and Hispanic, respectively. The percentage of male patients was slightly higher than that of female patients (53.9% versus 46.1%). Their average body mass index (BMI) was 29.3%. Most of the patients had other chronic health conditions, such as cardiovascular (88.2%), hypertension (76.4%), and kidney disease (65.6%).

**Table 1.**
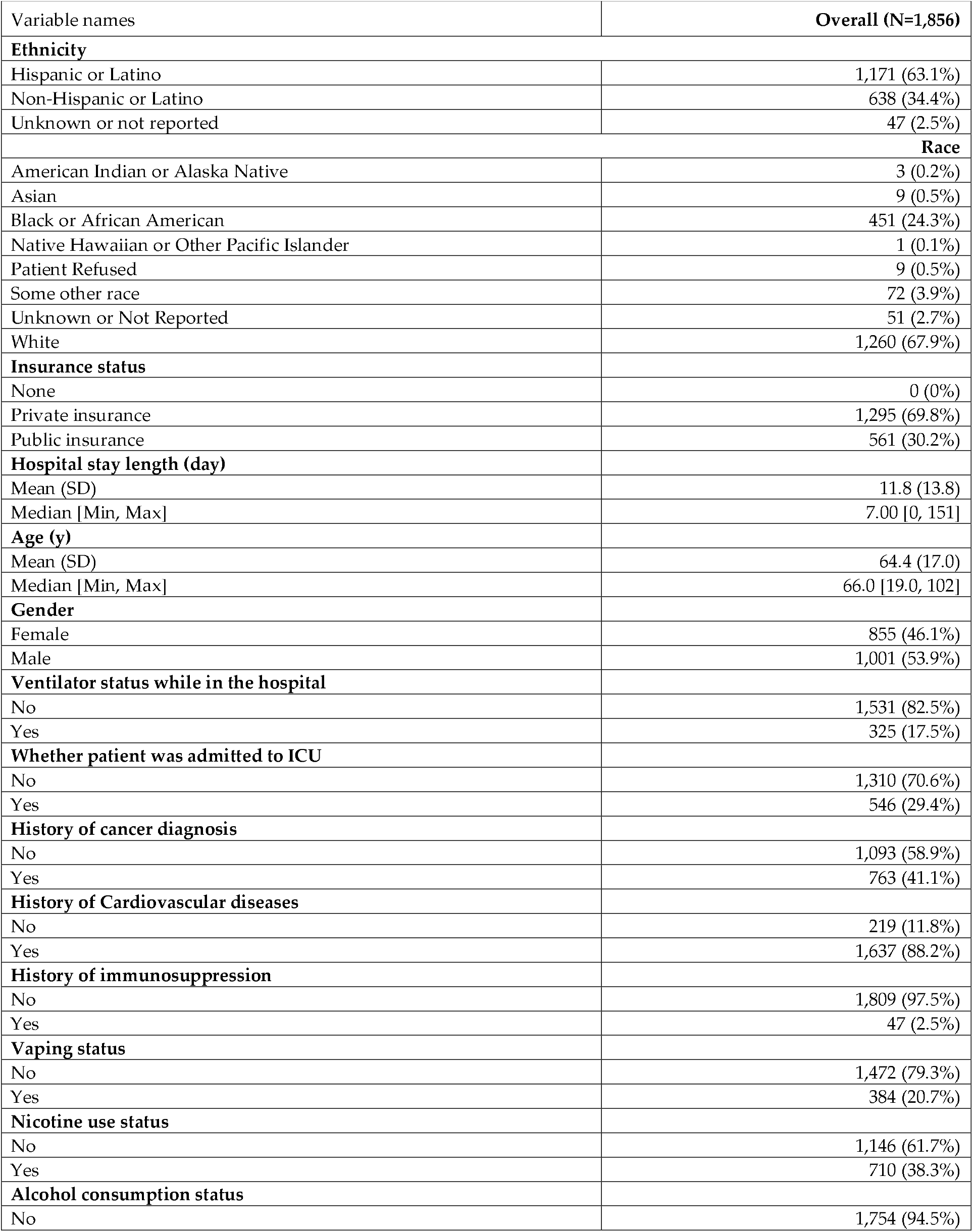
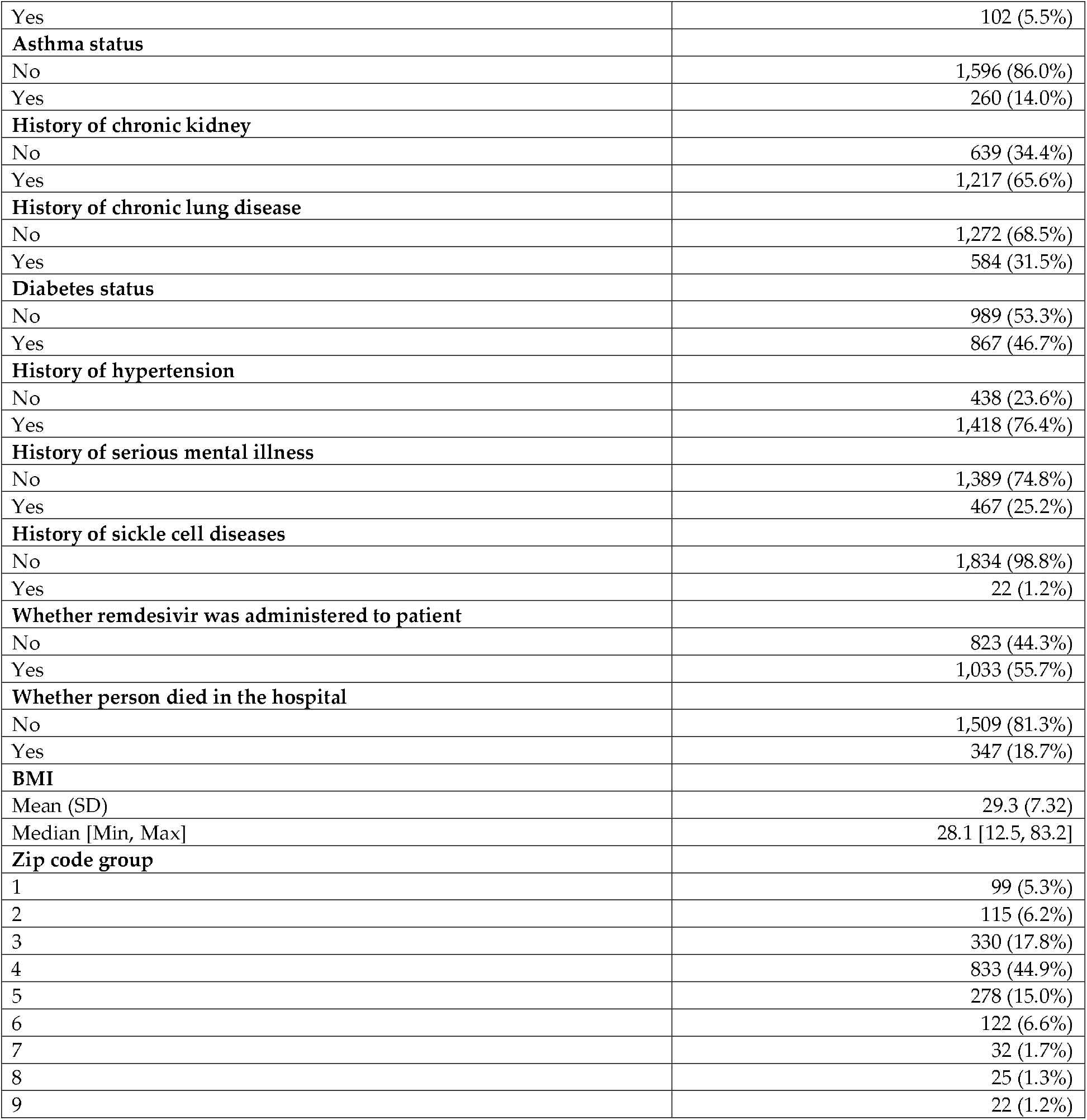
Summary statistics of patients’ characteristics.

Patients were stratified based on similarities in their socio-demographic characteristics, pre-existing health conditions, severity of COVID-19 infections, insurance status etc. Using factor analysis, these variables were collapsed into two components (Figure 1), which were then used for hierarchical clustering (of patients). Using the average silhouette, we chose the top four clusters. Patients’ characteristics are summarized by these four clusters in Table 2. The first cluster is dominated by preexisting chronic diseases. The second cluster is dominated by minority population; third cluster represents predominantly elderly white Hispanic population, and the fourth cluster represents the severity of COVID-19 infection and preexisting chronic diseases.

**Table 2.** Patients’ characteristics by four clusters.

| Characteristic | Cluster 1, N =334 | Cluster 2, N = 383 | Cluster 3, N = 786 | Cluster 4, N = 353 |
| --- | --- | --- | --- | --- |
| **Ethnicity** |  |  |  |  |
| Hispanic or Latino | 235 (70%) | 30 (7.8%) | 658 (84%) | 248 (70%) |
| Non-Hispanic or Latino | 83 (25%) | 351 (92%) | 112 (14%) | 92 (26%) |
| **Race** |  |  |  |  |
| Black or African American | 32 (9.6%) | 340 (89%) | 23 (2.9%) | 56 (16%) |
| White | 264 (79%) | 41 (11%) | 708 (90%) | 247 (70%) |
| **Insurance** |  |  |  |  |
| Private insurance | 313 (94%) | 297 (78%) | 451 (57%) | 234 (66%) |
| Public insurance | 21 (6.3%) | 86 (22%) | 335 (43%) | 119 (34%) |
| Patient's stay length (days) | 5 (4, 8) | 6 (4, 11) | 7 (4, 11) | 20 (12, 35) |
| Age (year) | 46 (37, 59) | 61 (48, 70) | 73 (64, 83) | 69 (59, 77) |
| Patient on ventilator | 7 (2.1%) | 8 (2.1%) | 4 (0.5%) | 306 (87%) |
| Patient in ICU | 46 (14%) | 56 (15%) | 101 (13%) | 343 (97%) |
| Cancer diagnosis | 90 (27%) | 179 (47%) | 370 (47%) | 124 (35%) |
| Cardiovascular disease | 132 (40%) | 378 (99%) | 786 (100%) | 341 (97%) |
| Vaping | 14 (4.2%) | 26 (6.8%) | 39 (5.0%) | 305 (86%) |
| Chronic kidney disease | 80 (24%) | 303 (79%) | 519 (66%) | 315 (89%) |
| Chronic lung disease | 42 (13%) | 153 (40%) | 267 (34%) | 122 (35%) |
| Diabetes | 33 (9.9%) | 235 (61%) | 396 (50%) | 203 (58%) |
| Hypertension | 36 (11%) | 354 (92%) | 746 (95%) | 282 (80%) |
| Serious mental illness | 25 (7.5%) | 115 (30%) | 244 (31%) | 83 (24%) |
| Administered Remdesivir | 202 (60%) | 198 (52%) | 378 (48%) | 255 (72%) |
| Patient mortality | 6 (1.8%) | 23 (6.0%) | 108 (14%) | 210 (59%) |
| <sup>1</sup> n (%); Median (IQR) |  |  |  |  |

Note: Cluster 1: Adult population with low prevalence of preexisting health conditions; Cluster 2: Elderly African American; Cluster 3: Elderly Hispanic white; Cluster 4: Severity of COVID-19 and pre-existing conditions.

**Figure 1.**
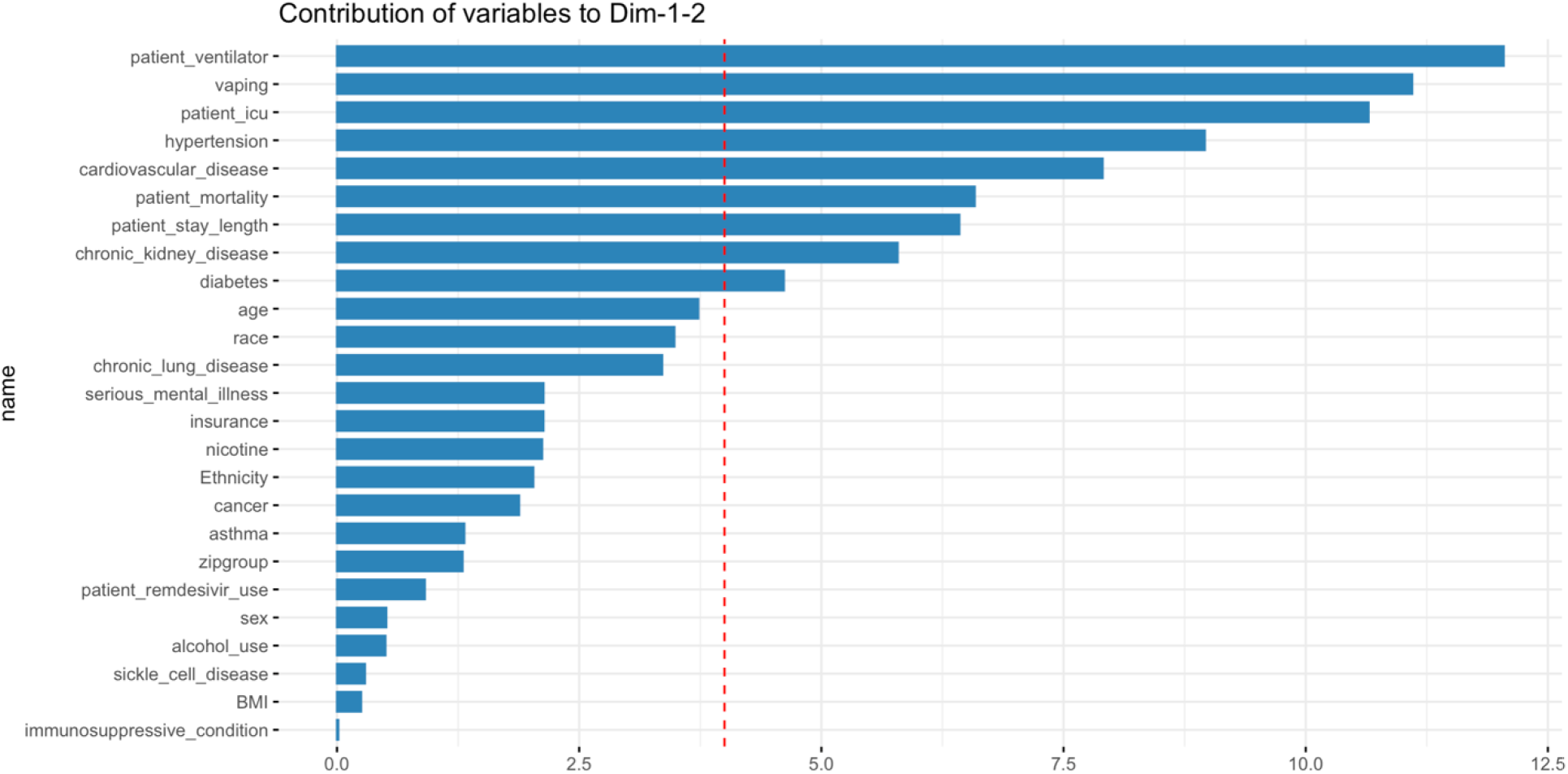
Contribution (%) of variables to the first 2 principal components.

The fatality rate among the admitted patients was 18.7% (or 347 of 1,856). The cluster 2 that represented predominantly white Hispanic patients with hypertension and cardiovascular diseases have the highest incidence of COVID-19 hospital admission (820 (44%)). However, the fatality rate due to COVID-19 was highest in cluster 4 that represented patients with multiple chronic health conditions. Cluster 1 that represented adult patients with less prevalence of chronic health conditions had the lowest COVID-19 related fatality rate (1.8%).

An exploratory analysis of cross-correlation suggested that there was a significant correlation between COVID-19 hospital admission and time-lagged WW-SARS-CoV-2 (Figure 2). The highest correlation of COVID-19 hospital admission was observed with the four-week lagged WW-SARS-CoV-2. Moreover, WW-SARS-CoV-2 data had strong temporal autocorrelation. Thus, we computed autocorrelation weighted average of WW-SARS-CoV-2, which represented all significant time-lagged WW-SARS-CoV-2. Given that the time-series COVID-19 data were also autocorrelated, we added a week-lag COVID-19 cases as an autoregressive term to account for the autocollinearity and it also served as a proxy of the transmission rate of the disease.

**Figure 2.**
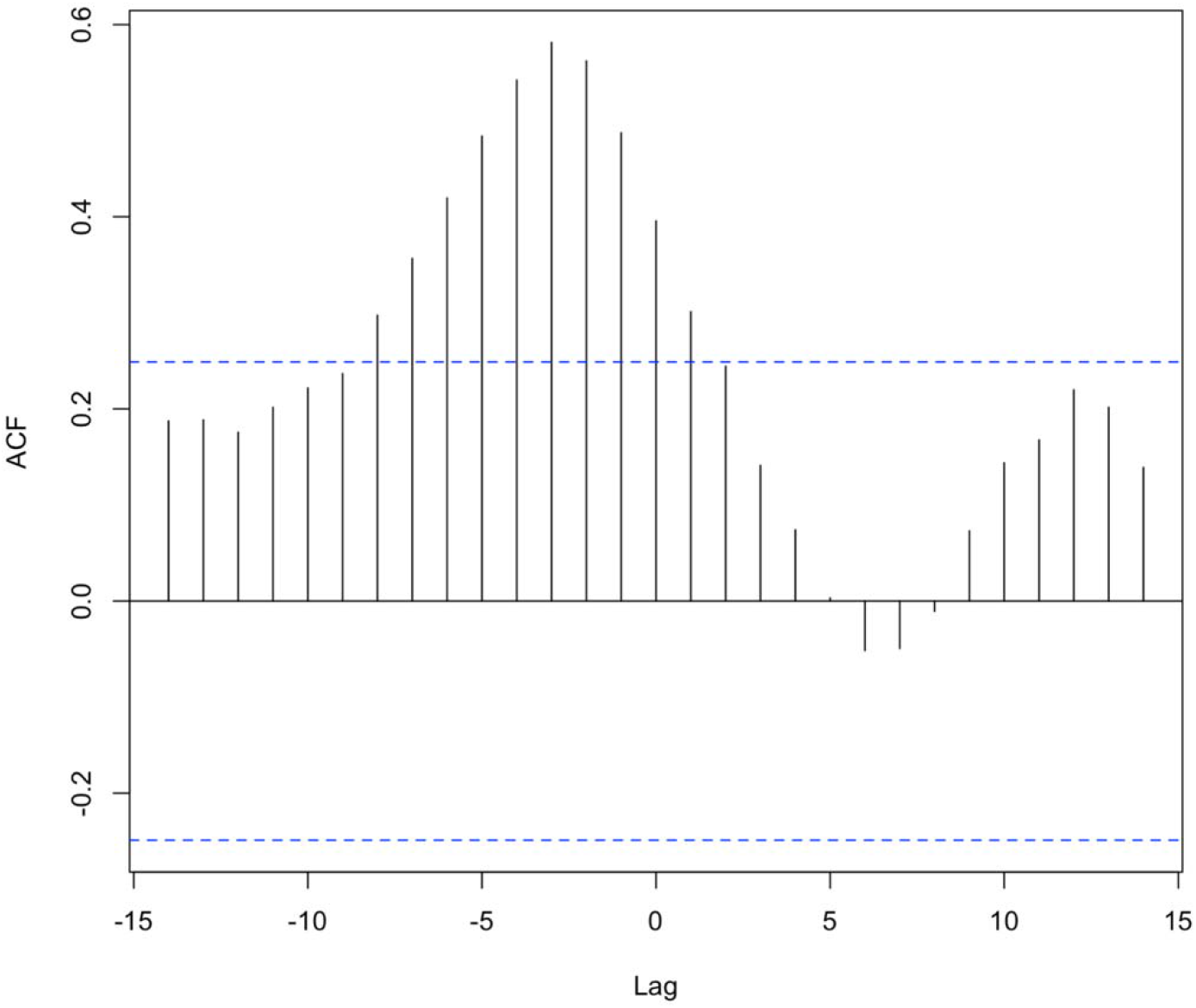
Cross-correlation between COVID-19 hospital admission and time-lagged WW-SARS-CoV-2.

Our results suggest that there was a significant relationship between COVID-19 hospital admission and the concentration of WW-SARS-CoV-2 (Table 3). One percent increase in the WW-SARS-CoV-2 was associated with an 0.28% increase in the number of COVID-19 cases admitted to the hospital (β ∼ 0.28; 95% CI = 0.18 to 0.37; p < 0.01). A week-lagged autoregressive term of COVID-19 cases also showed a strong association, e.g. 1% increase in a week lag COVID-19 cases was associated with a 0.005% increase in the risk of COVID-19 hospital admission, suggesting a rapid transmission of the disease and its severity. Both WW-SARS-CoV-2 and a week lag COVID-19 hospital admission together explained 89% of the variations in the COVID-19 related hospital admission.

**Table 3.** Association between weekly COVID-19 hospital admission and WW-SARS-CoV-2.

|  | Estimate (95% CI) |
| --- | --- |
| 1-4 week lag autocorrelation weighted $\log_{10}(\text{WW-SARS-CoV-2 (GC/L)})$ | 0.084 **<br>(0.04, 0.13) |
| $\log_{10}(\text{1 week lag COVID-19 cases})$ | 0.76 ***<br>(0.66, 0.86) |
| Adjusted R<sup>2</sup> | 0.93 |
|  | -96.85 |
| *** p<0.01, ** p<0.05, * p<0.1; 95% confidence interval in parentheses. |  |

The association between WW-SARS-CoV-2 and COVID-19 hospital admission varied across four clusters. This association was strongest in cluster 2 followed by cluster 1 and 3 (Table 4). In cluster 2 that predominantly represented minority communities, the association between COVID-19 hospital admission and WW-SARS-CoV-2 was the strongest (β ∼ 0.21; 95% CI = 0.06 to 0.37; p < 0.01). However, in cluster 4 that represented more severe cases of COVID-19 infections the association between COVID-19 cases and WW-SARS-CoV-2 was insignificant. In this cluster 87% of the patients were on ventilator and 98% of them had to be admitted to ICU. But in this cluster, a week-lag autoregressive term of COVID-19 cases showed the strongest association with the hospital admission of more serious cases of COVID-19; 1% increase in the hospital admission of severe cases of COVID-19 infection a week before was indicator of a 0.03% increase in hospital admission of such type of cases a week after (β ∼ 0.029; 95% CI = 0.2 to 0.037; p < 0.01).

**Table 4.**
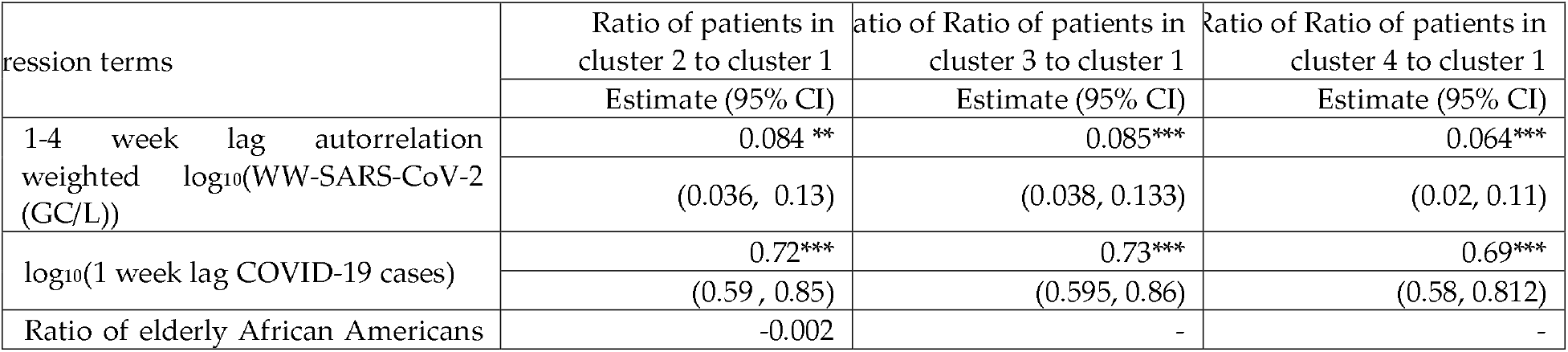
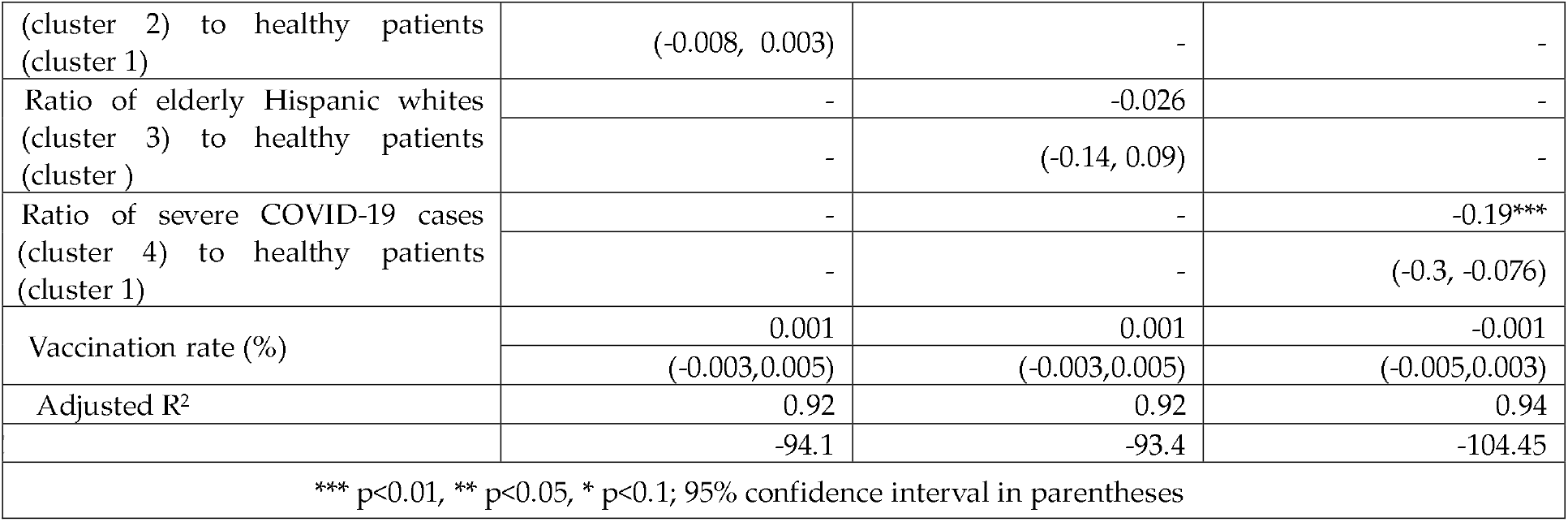
Association between weekly COVID-19 hospital admission and WW-SARS-CoV-2 across four groups of patients.

Mortality among COVID-19 cases admitted to the hospital did not show a significant association with the WW-SARS-CoV-2 (Table 5). However, the autoregressive term of a week-lag mortality and county level vaccination showed significant associations with COVID-19 mortality. With 1% increase in vaccination rate (in the Miami-Dade County) the COVID-19 mortality risk in patients admitted to the UM clinics declined by 0.05% (β ∼ -0.048; 95% CI = -0.062 to -0.034; p < 0.01), and 1% increase in one-week lag mortality indicated a 0.05% higher risk of COVID-19 mortality in patients admitted to UM clinics.

The relation between COVID-19 mortality, WW-SARS-CoV-2 and vaccination rate varied across four clusters (or groups) of patients. The time lagged WW-SARS-CoV-2 showed a significant inverse association with COVID-19 mortality for clusters 2 and 4 (Table 6), which is counter-intuitive. The vaccination was most effective in cluster 4 and cluster 3; with 1% increase in vaccination rate the risk of COVID-19 mortality declined by 0.024% for cluster 4 (which represented severe cases of COVID-19 infection with preexisting health condition) and 0.02% for cluster 3, (which represented White Hispanic population) (β ∼ -0.024; 95% CI = -0.034 to - 0.013; p < 0.01; β ∼ -0.02; 95% CI = -0.031 to -0.01; p < 0.01, respectively). However, vaccination did not show a significant association with COVID-19 mortality in cluster 1 and cluster 2, which represented patients with preexisting health conditions and minority populations, respectively.

**Table 6.** Association between weekly COVID-19 related mortality in patients admitted to UM hospitals and WW-SARS-CoV-2 and vaccination rate adjusted for proportion of patients in different clusters.

| Regression terms | Ratio of patients in cluster 2 to cluster 1 | Ratio of Ratio of patients in cluster 3 to cluster 1 | Ratio of Ratio of patients in cluster 4 to cluster 1 |
| --- | --- | --- | --- |
|  | Estimate (95% CI) | Estimate (95% CI) | Estimate (95% CI) |
| 1-4 week lag autorrelation weighted log<sub>10</sub>(WW-SARS-CoV-2 (GC/L)) | 0.0096 | -0.004 | 0.04 |
|  | (-0.13, 0.15) | (-0.14,0.13) | (-0.099, 0.176) |
| log<sub>10</sub>(1 week lag of COVID-19 mortality) | 0.72*** | 0.7*** | 0.71*** |
|  | (0.51, 0.93) | (0.49, 0.92) | (0.5, 0.915) |
| Ratio of elderly african americans to healthy patients | 0.0004 | - | - |
|  | (-0.016, 0.016) | - | - |
| Ratio of elderly hispanic whites to healthy patients | - | -0.004 | - |
|  | - | (-0.014, 0.007) | - |
| Ratio of severely affected to healthy patients | - | - | 0.006 |
|  | - | - | (-0.003, 0.015) |
| Vaccination rate (%) | -0.024** | -0.03** | -0.022** |
|  | (-0.043, -0.004) | (-0.046, -0.006) | (-0.04, -0.004) |
| Adjusted R<sup>2</sup> | 0.9 | 0.9 | 0.9 |
|  | 19.02 | 18.5 | 17.1 |
| *** p<0.01, ** p<0.05, * p<0.1; 95% confidence interval in parentheses |  |  |  |

## 4. DISCUSSION

Using the COVID-19 hospital admission data with detailed patient-specific characteristics and WW-SARS-CoV-2 data from a controlled hospital setting, this study demonstrates that there was a statistically significant relationship between COVID-19 hospital admission and WW-SARS-CoV-2, but this relationship varied by patients’ socio-demographics, preexisting health conditions and severity of the infection among patients. Although WW-SARS-CoV-2 did not show any significant association with COVID-19 mortality, the risk of morality decreased significantly with the increase in (COVID-19) vaccination rate.

This paper makes novel contributions. First, this paper documents the empirical association between time-lagged WW-SARS-CoV-2 and COVID-19 hospital admission and COVID-19 mortality from a controlled hospital setting. Hospital admission and WW-SARS-CoV-2 data used in the paper came from a hospital. Thus, SARS-CoV-2 in the wastewater represented patients in the hospital. Hospital staff could have shed the virus if they were infected. However, hospital staff are unlikely to be COVID-19 positive, given higher testing, vaccination quarantine standards for them. Although many studies document positive association between COVID-19 hospitalization and WW-SARS-CoV-2, the WW-SARS-CoV-2 data used in these other studies were from the city or municipality wastewater treatment plant(s), which might have also included SARS-CoV-2 shed by infected and/or asymptomatic individuals who were not admitted to the hospital.^25,26^ The overall findings of the association between WW-SARS-CoV-2 COVID-19 cases is consistent with the recent literature,^1,27,28^ but the strength of this relationship is stronger than that documented in the literature. Second, this paper documents variations in the relationship between time-lagged WW-SARS-CoV-2 and COVID-19 hospitalization by patient specific characteristics.

There is overwhelming body of literature that documents age, gender, income and race disparities in the risks of contracting COVID-19, hospitalization and COVID-19 mortality, ^9,20,29,30^ e.g. a study reported that African Americans were 2.7 times more likely to be hospitalized for COVID-19 than other groups.^29^ However, this is the first attempt that documents variations in the relationship between COVID-19 hospital admission and WW-SARS-CoV-2 by patient specific characteristics. For example, the relationship between WW-SARS-CoV-2 and COVID-19 cases was insignificant in Cluster 4, which represented serious cases of COVID-19. This suggests that when patients are intubated or in the ICU, they are incapacitated and may not be shedding the virus in the bathroom, which will result in low concentration of WW-SARS-CoV-2 even though the number of serious patients admitted to the hospital can be high.

The findings of the paper have implications for developing and validating infectious diseases surveillance and management of the wastewater from hospitals. Like SARS-CoV-2, other pathogens, such as *Candida auris*, monkey pox and polio, have been traced in the wastewater samples.^31-36^ Thus, proactive monitoring of pathogens in wastewater offers an unprecedented opportunity for infectious diseases surveillance. The findings presented in this paper suggest that the effectiveness of infectious disease surveillance using wastewater can be greatly affected by patients’ socio-demographic characteristics and preexisting health conditions. Thus, developing effective surveillance will require deep understanding of the time-space dynamic of the targeted pathogen and its testing and validation in diverse communities. The wastewater from the hospitals serving infected individuals will have elevated levels of SARS-CoV-2. Thus, this may contaminate the environment if it is released untreated because pathogens, including SARS-CoV-2, degrade slowly.^31,37^ Thus, there is a need for treating the wastewater from such facilities before it released to the main sewer system.^38,39^

Despite the scientific contributions of this paper, the findings reported in the paper must be interpreted in the light of its limitations. First, the efficacy of SARS-CoV-2 in the wastewater samples to predict COVID-19 cases and mortality can be influenced by the reliability of virus recovery from the wastewater samples. For example, the detection and recovery of the virus in the sample can be subject to the physical and chemical composition of the sample and processing error. Second, the mismatch in the temporal (and spatial) resolution of health and environmental data can influence model inferences. For example, COVID-19 hospital data were reported daily, and wastewater samples were collected weekly. Thus, matching both data sets by exact day resulted in reduced sample size and interpolating weekly data by every day introduced uncertainty. Finally, the findings of this study from a control hospital setting constrain the generalizability of the study to a community setting. Despite these limitations, the findings of the paper have potential to guide effective infectious risk surveillance based on the community specific characteristics and health conditions.

## 5. CONCLUSIONS

Our results indicate that the COVID-19 hospital admission and mortality varied by patients’ characteristics, and the association of WW-SARS-CoV-2 and COVID-19 hospital admission also varied by patients’ socio-demographics and preexisting health conditions. Thus, the efficacy COVID-19 surveillance using WW-SARS-CoV-2 can be improved by adjusting for patients’ demographics and preexisting health conditions. These findings can also be extrapolated for the surveillance of other infectious diseases.

## Data Availability

All time-stamped wastewater SARS-CoV-2 and aggregated patients data can be provided upon request to the corresponding author.

## ACKNOWLEDGEMENTS

This work in part was supported by NIH (U01DA053941). All clinical data (COVID-19 case and mortality rates for UM hospitals) are confidential data. However, the weekly SARS-CoV-2 concentration in the wastewater samples can be provided upon a reasonable request to the corresponding author.

